# Predictive usefulness of PCR testing in different patterns of Covid-19 symptomatology - Analysis of a French cohort of 12,810 outpatients

**DOI:** 10.1101/2020.06.07.20124438

**Authors:** AP-HP / Universities / Inserm COVID-19 research collaboration members, Apra Caroline, Caucheteux Charlotte, Mensch Arthur, Mansour Jenny, Bernaux Mélodie, Dechartres Agnès, Debuc Erwan, Lescure Xavier, Dinh Aurélien, Yordanov Youri, Jourdain Patrick, Mensch Arthur, Caucheteux Charlotte, Apra Caroline, Mansour Jenny, Paris Nicolas, Gramfort Alexandre

## Abstract

Polymerase Chain reaction (PCR) is a key tool to diagnose Covid-19. Yet access to PCR is often limited. In this paper, we develop a clinical strategy for prescribing PCR to patients based on data from COVIDOM, a French cohort of 54,000 patients with clinically suspected Covid-19 including 12,810 patients tested by PCR. Using a machine learning algorithm (a decision tree), we show that symptoms alone are sufficient to predict PCR outcome with a mean average precision of 86%. We identify combinations of symptoms that are predictive of PCR positivity (90% for anosmia/ageusia) or negativity (only 30% of PCR+ for a subgroup with cardiopulmonary symptoms): in both cases, PCR provides little added diagnostic value. We deduce a prescribing strategy based on clinical presentation that can improve the global efficiency of PCR testing.

## Introduction

The coronavirus disease (Covid-19) epidemic started in China in December 2019 and has spread worldwide, infecting more than 5.5 million people by May 25th (1). Making the diagnosis of Covid-19 infection can be difficult, since the clinical presentation is versatile, including associations of fever, myalgia, fatigue, cough, shortness of breath, gastro-intestinal signs, headaches, upper respiratory tract symptoms… (2). Several tests have proved helpful to diagnose Covid-19 infection (3–5). The current reference test is reverse transcriptase polymerase chain reaction (PCR), which attests the presence of viral RNA in the sample, usually nasopharyngeal swabs (6).

There is a strong need for a practical strategy to approach diagnostic investigations. PCR is costly and remains difficult to use in practice with a significant time from sampling to results (7). Moreover, PCR is highly specific of the presence of viral nucleic acid in the sample, yet it lacks sensitivity: a negative test does not negate the possibility that an individual is infected (4). This creates a diagnostic doubt, and first-line alternative investigations, such as chest imaging, may sometimes be more relevant.

We analyze the PCR usefulness as a diagnostic tool in different clinical presentations, in order to develop and assess a strategy for PCR prioritization in patients with suspected Covid-19. For this, we first provide a statistical description of the PCR positive population; we use propensity-score weighting to correct for the non-homogeneity of testing in the whole cohort. We then identify combinations of symptoms that are predictive of PCR result, using decision trees, in a supervised machine learning approach. We base our analyses on a large ambulatory cohort of 54,000 patients followed by an unique telemonitoring platform in the greater Paris region.

## Methods

We first describe the access to PCR testing, based on patients’ characteristics and symptoms. We study whether PCR-positive (PCR+) and PCR-negative (PCR-) patients have different clinical profiles. We then perform a multivariate predictive study: we identify combinations of symptoms that are predictive of either high or low chance of PCR positivity with weighting on the propensity score for PCR testing. Based on these identified combinations, we propose a triage strategy to target PCR testing in patients for whom PCR results will bring the highest additional information for Covid-19 diagnosis.

### Population - COVIDOM telemonitoring program

In France, a telemonitoring web-application called COVIDOM has been developed for home management of suspected or confirmed Covid-19 patients. In this application, self-administered daily questionnaires can trigger alerts that are handled in a regional medicalized control center. It was launched in the Greater Paris area on March 9th and aims at efficiently detecting patients at risk of deterioration while relieving the burden for healthcare professionals. Patients are included in COVIDOM after seeking medical care in an outpatient setting (emergency services or general practitioners) or after being discharged from hospital. We excluded all patients under 18.

### Data

Patients in COVIDOM filled out questionnaires, specifying characteristics (age, sex, weight, height), comorbidities (diabetes, hypertension, chronic obstructive pulmonary disease, asthma, heart failure, coronary heart disease, cancer under treatment, chronic kidney disease, other chronic disease), smoking status, symptoms since the beginning of the suspected Covid-19 disease (fatigue, myalgia, breathlessness, ageusia, anosmia, anorexia, chest pain, chest oppression, cough, fever, diarrhea, vomiting, shivers, rash, frostbites, conjunctivitis, other symptoms), hospitalisation history, investigations that were performed (PCR, chest CT-scan, chest X-ray), and PCR results. The questionnaire is available in Fig.S1. These patient-reported data are completed with PCR results from the *French Assistance Publique - Hôpitaux de Paris* (AP-HP) data warehouse, also known as *Entrepôt de Données de Santé* (EDS). AP-HP is the network of all university hospitals in the greater Paris region. PCR were performed according to international guidelines on respiratory samples, mainly nasopharyngeal swabs (6).

During registration, patients provided an electronic consent for the Covidom telemonitoring program and they were informed of the potential use of anonymized data for research purposes. This use was approved by the Scientific and ethical committee of APHP (IRB00011591).

### Analyses

#### Access to PCR testing

In a retrospective analysis, we seek to identify patients who had PCR testing (Fig.1), and to report associations of patient characteristics with PCR results (Fig.2). For this, we consider the following covariates: sex; age (quantized in 5 groups); tobacco consumption (current smoker or not); comorbidites: respiratory, cardio-vascular, diabetes or obesity; presence of symptoms: breathlessness, anorexia, tiredness, digestive signs (diarrhea or vomiting), conjunctivitis, cutaneous symptoms (rash or frostbites), shivers, myalgia, cough, fever, cardiopulmonary symptoms (breathlessness associated to chest pain or chest oppression) or chemosensory impairment (anosmia or ageusia); ambulatory status (has the patient been hospitalized or not).

**Figure 1:**
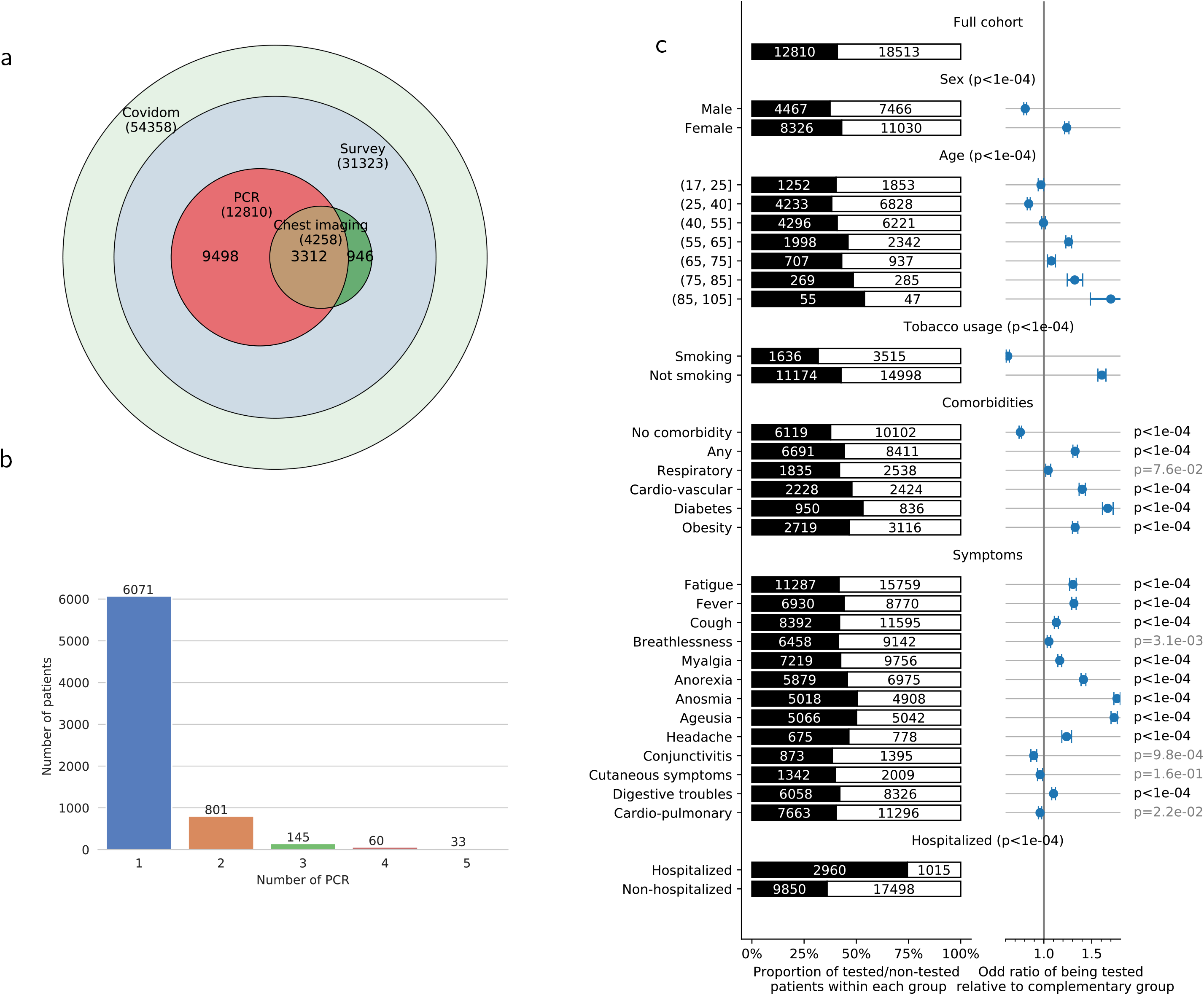
Differentiated access to PCR testing in patients who answered the COVIDOM survey (N=31,323). a) Description of the investigations in the COVIDOM cohort combining the survey and the EDS database: 54,358 patients are included in the web-application for daily monitoring, among which 31,323 answered the complete survey. b) Description of repeated PCR testing for patients included in the Corona OMOP database (N=6,621). Patients benefited from 1 to 10 PCR tests each. Median time between PCR1 and PCR2 was 8 days, PCR1 and PCR3: 13 days, PCR1 and PCR4: 15 days, PCR1 and PCR5: 21 days. c) Access to PCR testing in patients who answered the COVIDOM survey, as a function of various patient characteristics. The size of the black bar indicates the proportion that has been tested of a given group.. The population is stratified based on demographic characteristics, tobacco usage, comorbidities (“any” includes any of the following or hypertension, chronic kidney disease, cancer under treatment or other as indicated by the patient, “respiratory” indicates asthma or COPD, “cardio-vascular” indicates heart failure or coronary disease, obesity a BMI above 30), symptoms experienced at some point of the disease (“cardiopulmonary” indicates breathlessness associated to chest oppression or chest pain), need for admission in hospital before or after inclusion in COVIDOM. The right column indicates the odd-ratios of being tested in each group, compared to the complementary group. We test whether these odds ratios significantly differ from 1 using a Wald test. Table.S1 provides all numerical data.

**Figure 2:**
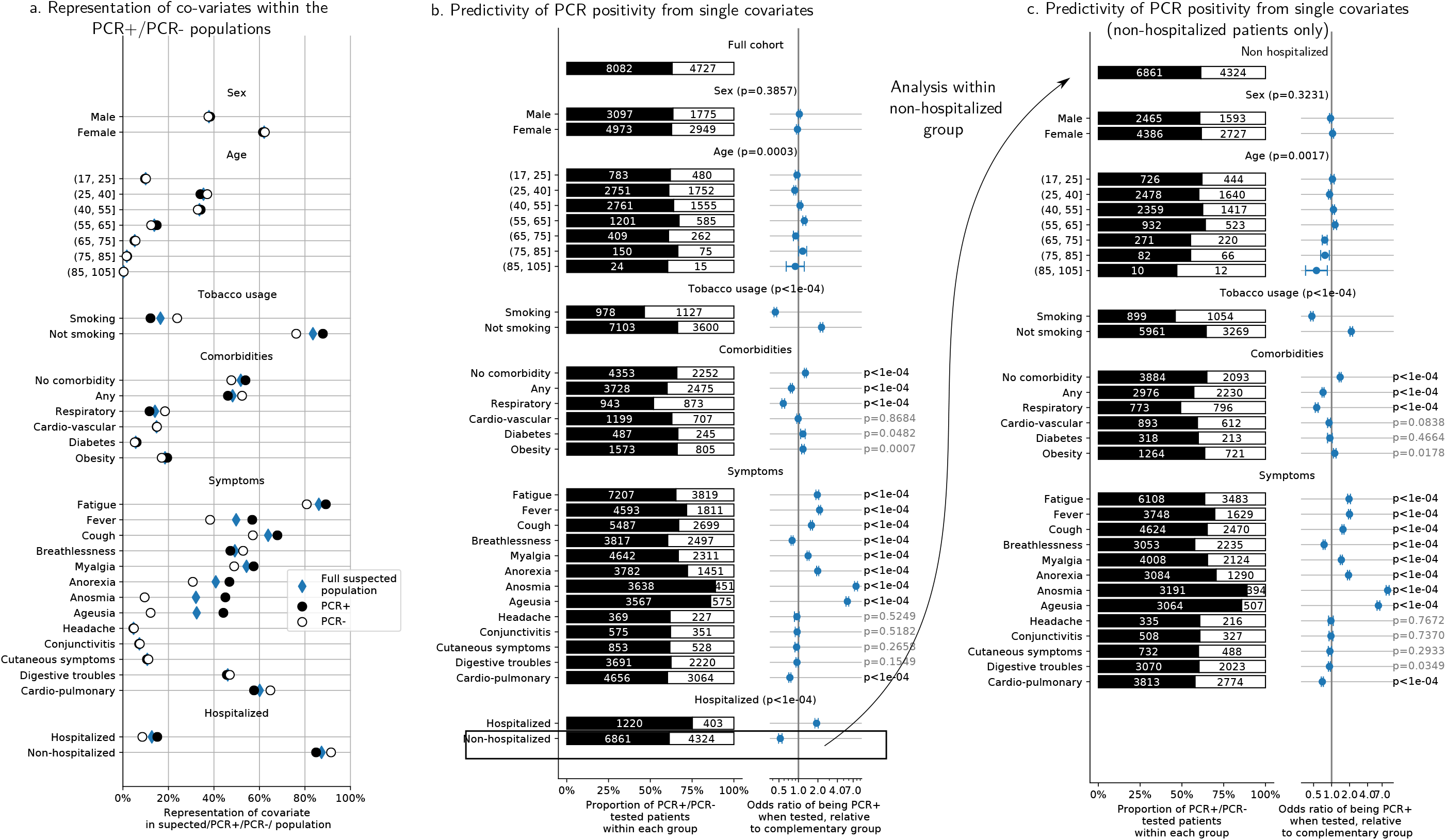
Effects of patient characteristics on PCR results. We apply a propensity-scoring weight to each patient, to remove the testing-propensity confound. We report weighted counts/proportions/odds-ratios. a. Representation of each characteristic in the whole cohort (blue diamonds), the PCR+ cohort (black dot) and the PCR-cohort (white dot). b. PCR results as a function of different characteristics in the tested population (cf Fig. 1c for categories). Odds ratios of being PCR+ when belonging to a given subgroup, and p-values that these ratios are significantly different from 1 (using a Wald test). Patients without comorbidities nor smoking are more likely to be PCR+, so do patients with anosmia, ageusia, anorexia, fever, fatigue, cough, myalgia. On the opposite, comorbidities, in particular respiratory diseases, and symptoms such as breathlessness and cardiopulmonary symptoms are associated with PCR-. c. Same analysis as b, within the subpopulation that has not been hospitalized. Results are similar. Table.S1 provides all numerical data.

Within the population of patients who answered the questionnaire, we evaluate the associations between patient characteristics and PCR testing: for this, we estimate odds ratio for each variable using univariate logistic regression models. We test whether the odds ratio significantly differs from 1 using a Wald test.

#### Associations of patient characteristics with PCR results

Within the tested population, we evaluate the association of each covariate with PCR results. To account for the lack of homogeneity in testing and correct for a possible indication bias, we use a propensity score for weighting each patient: for this, we estimate the probability of being tested using a multivariate logistic model (with all covariates introduced above). Weighting results in a pseudo-population of size twice the number of tested patients, where covariates proportion are similar within the whole cohort and the tested population.

We then estimate one univariate logistic regression model per covariate to predict PCR positivity, and report associated pseudo-counts (i.e. counts weighted using the propensity score) and odds ratios. Complementarily, we compare the weighted proportion of a given covariate within the PCR+ and PCR-populations.

#### Predicting PCR results from patient symptoms

To prioritize patients due for a PCR test, we seek combinations of symptoms that are predictive of the PCR result. For this, we estimate a multivariate decision tree (8) that, for each patient, predicts the result of the PCR test based on his/her symptoms. A decision tree recursively splits the population based on the presence or not of a given symptom, so as to progressively separate PCR+ and PCR-into different groups. It thus automatically provides predictive combinations of symptoms, whose importance is then verified on a set of patients not used for estimation. As in the univariate analysis, we use propensity score weighting during estimation and evaluation.

We train a decision tree on 80% of the tested patients and evaluate its performance on the 20% held-out group. We repeat the training procedure across multiple separations of training and held-out data to evaluate the variance of the predictive model performance. Details on decision-tree parameters, architecture choices and weighting procedure are reported in the supplementary material. We report precision-recall curves, average precision of the model, and a description of the splits performed by the trained decision tree. We evaluate the importance of each symptom in predicting the PCR outcome by measuring how hiding this observed variable from the decision tree affects its performance (9).

Finally, in each group defined by the decision tree, we report the odds ratio of being PCR+, and report PCR+ proportion. Odds and proportions are weighted by propensity scores. We simplify the decision tree to propose actionable rules to prioritize PCR access.

## Results

### Cohort description

From inception to May 6th, 54,358 patients were registered in COVIDOM by a physician for daily monitoring, 31,323 answered the questionnaire (flow-chart of Fig.1a). 3774 patients (12%) were included after hospitalization. There was a median of 16 days (IQ9-23) after the first symptoms and 10 days (IQ2-16) after the inclusion in COVIDOM when the patients filled up the forms (Fig.S2).

The mean age of the patients is 43.6 (SD14.3) with 28779 (92%) under 65 year-old. As detailed in Fig.2 and Table S1, the most frequent symptoms in the whole cohort were fatigue (86%), cough (64%), myalgia (54%), fever (50%), breathlessness (50%), and digestive symptoms (46%). Breathlessness associated with chest oppression or pain was mentioned by 61%. Anosmia and ageusia are present in respectively 32% and 32% of patients, with 26% presenting both symptoms. Anosmia or ageusia is more frequent in women (28% of women present both symptoms versus 22% of men, p<0.0001), and the mean age of patients with chemosensory impairment is 42.2 years (SD13.2), younger than the rest of the cohort (p<0.0001).

In total, 12,810 patients (41%) were tested by PCR, after we excluded 75 patients with undetermined results (0.6%). Chest imaging was performed in 5,010 patients(16%). In patients who had PCR, the mean number of PCR was 1.2+/-0.6 and the median time between PCR1 and PCR2 was 7 days (IQ2-19) (Fig.1b and Fig.S3).

### Differentiated access to PCR testing in the COVIDOM cohort

Studying PCR access in the COVIDOM cohort shows that the test is not systematically performed for all symptomatic patients, as detailed in Fig.1c and Fig.S5. Patients more prone to be tested are women (43% vs 37%, p<0.0001), elderly patients (p<0.0001), and non-smokers (43% vs 32%, p<0.0001). Patients with comorbidities are tested more often (44% vs 37%, p<0.0001), especially patients with diabetes (53%), cardio-vascular disease (48%) or obesity (47%), but not patients with respiratory comorbidities. Concerning clinical presentation, patients with anosmia or ageusia are more likely to be tested (respectively 51% and 50%). On the opposite, patients with cardiopulmonary signs, i.e. breathlessness associated with chest oppression or chest pain, are as likely to be tested as the whole cohort (40%). As expected, patients who were hospitalized before or after their inclusion in COVIDOM were tested more often than outpatients (3,134, 80%, against 10,724, 40%).

### Associations of patient characteristics with PCR results

The remaining analyses are performed with propensity-score weighting; from now, we report counts, proportions and odds ratios for the weighted population, unless specified otherwise. Fig.S5 reports weighted counts and proportions in access to PCR testing: weighting ensures that the characteristics of the tested population are similar to those of the whole cohort.

PCR is positive in 63% of tested cases. We report results on the tested population in Fig.2a and 2b, and Table S1. We do not find any significant effect of age and sex. Tested smokers are less likely to be PCR+ (46% are PCR+ vs 66%, p<0.0001). Patients without comorbidities are more likely to be PCR+ (66% vs 60%, p<0.0001), as well as patients with anosmia, ageusia, anorexia, fever, fatigue, cough, myalgia (p<0.0001). In contrast, patients with breathlessness and cardiopulmonary symptoms are less likely to be PCR+ (60% vs 63%, p<0.0001), suggesting that PCR is less sensitive for patients with pulmonary symptoms than for other patients with suspected Covid-19. In echo to this observation, patients with respiratory comorbidities are less likely to be tested positive than other patients (52% vs 63%, p<0.0001). Other comorbidities have no significant association with PCR results.

Hospitalized patients are more likely to be PCR+ than non-hospitalized patients (75% vs 61%, p<0.0001), a potential cause of bias in our analysis; yet, as indicated in Fig.2c and Fig.S4, the findings above also hold within the population of non-hospitalized patients. We note that PCR tests performed more than 12 days after the first symptoms were 58% negative (4,2 times more negative than average, Fig.S3b).

### Anosmia/ageusia, cardiopulmonary signs and fever predict PCR result in patients with clinically suspected Covid-19 infection

The strong association between symptoms and PCR result encourages us to verify how symptoms effectively predict the PCR result. We focus on symptoms as predictive factors to train a decision tree for PCR testing.

On the held-out group (2,562 patients), the decision tree (trained on 10,248 patients) achieves 0.83 mean average precision (Fig.3c, 0.63 chance level). It identifies combinations of symptoms that efficiently separate PCR+ from PCR-patients (Fig.3a), and predict PCR results on newly seen patients. Permutation importance (PI, Fig.3b) tests show that anosmia/ageusia is the most important splitting criteria (PI=0.133 +/- 0.009), followed by cardiopulmonary symptoms (PI=0.017 +/- 0.004) and fever (PI=0.016 +/- 0.004). As reported in Fig.3a, in the evaluation cohort, 86% of the patients with anosmia/ageusia are PCR+ (OR 6.18, IC[5.89-6.47]). In the non anosmic/ageusic group (1,403 patients), patients with fever are less likely to be PCR+ (OR 0.83, IC[0.80-0.86]). Patients with cardiopulmonary symptoms and no fever are very unlikely to be PCR+ (OR 0.18, IC[0.17-0.20]). The decision tree splits on the held-out data and on the whole cohort are reported in Fig.S6 and Fig.S7, with associated values reported in tables S2 and S3. Overall, the trained decision tree identifies combinations of symptoms that are predictive of high chance of PCR positivation (anosmia or ageusia), or low chance of PCR positivation (no anosmia or ageusia, no fever but cardiopulmonary symptoms). Those respectively correspond to cases where Covid disease is very likely, and cases for which PCR has a low sensitivity. For patients experiencing such symptoms, performing a PCR has a low marginal value to adjust Covid-19 diagnosis.

**Figure 3:**
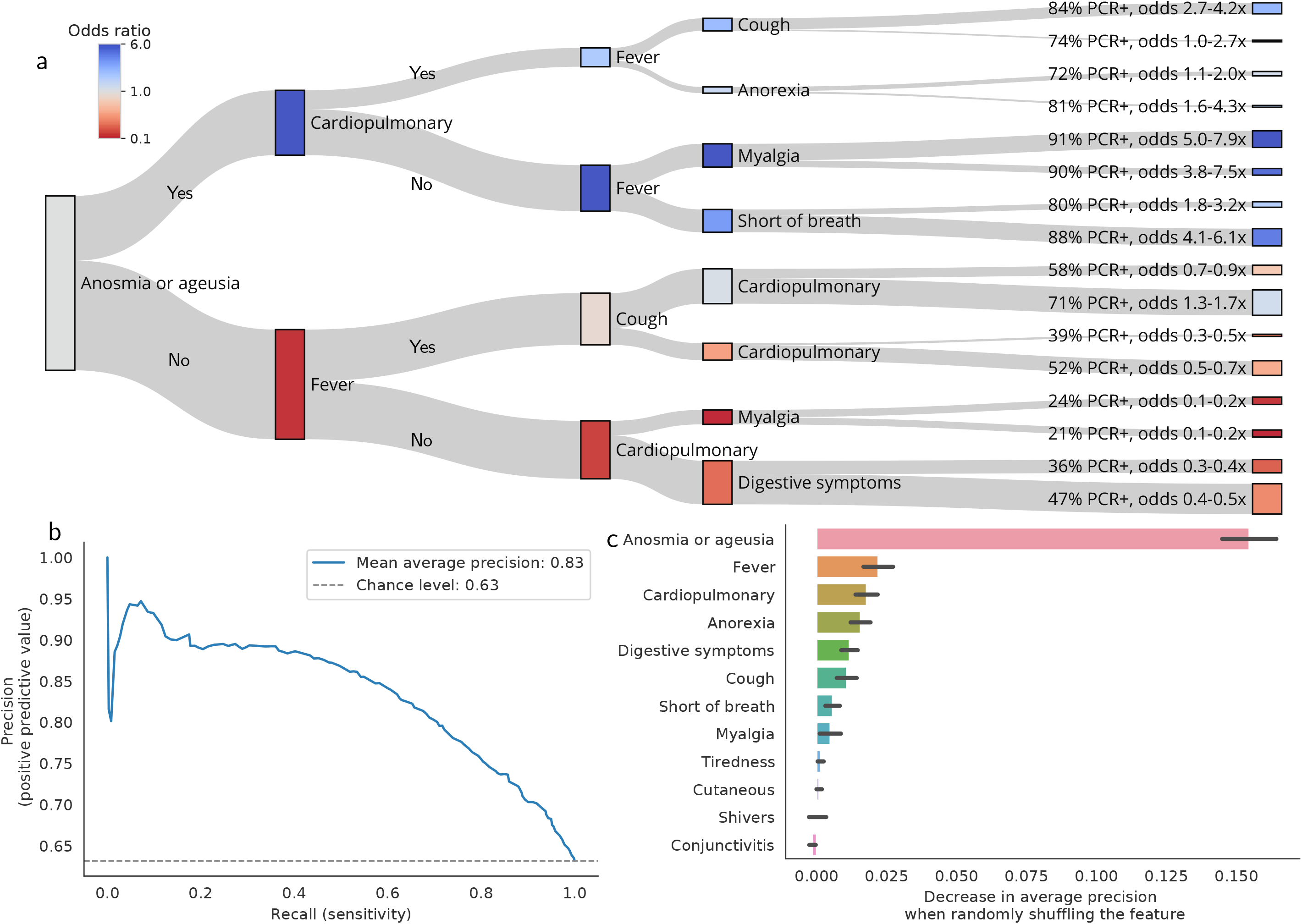
Marginal value of PCR testing in patients with clinically suspected Covid-19 infection. Estimation and evaluation using testing propensity-score weights. *a) Decision paths of the tree, applied to evaluation patients*. A decision tree classifier is trained on 80% of the tested patients. It predict PCR positivity using 12 features: breathlessness, anorexia, tiredness, digestive signs (diarrhea/vomiting), conjunctivitis, cutaneous symptoms (rash/frostbites), shivers, myalgia, cough, fever, cardiopulmonary symptoms (breathlessness + chest pain/oppression) and chemosensory impairment (anosmia/ageusia). We evaluate the decision tree on 20% held-out patients and illustrate how it splits this population. Each node is a splitting criterion (presence of the symptoms to the top, absence to the bottom). The colour of each node corresponds to the odds ratio of being PCR+ at this stage of the decision path. For each leaf, the probability and odds ratio of being PCR+ are reported (see Fig.S6, Fig.S.8 and Table.S1 for details). b) *Permutation features importance on the evaluation set*. The permutation importance is an indicator of the relevance of a feature at predicting PCR positivity. It measures the decrease in the model score (here, average precision) when a single feature is randomly shuffled. We report the permutation importance on the left-out evaluation data (20% of the dataset) for each feature of the decision tree. Error bars are the standard deviations of the importance through 50 different permutations. c) *Performance of the decision tree on the test set*. Precision-recall curve of the decision tree on the 20% held-out set of tested patients.

The findings that we report hold for multiple training/held-out data separation (Fig.S8); they remain similar without propensity score weighting (Fig.S9), and when performing the analysis within the population of ambulatory patients only (Fig.S10).

### Diagnostic strategy based on symptoms to target PCR testing in patients suspected with Covid-19 infection

We adapt the estimated decision tree into an actionable testing strategy based on clinical signs (Fig.4a), taking into account that PCR positivity establishes the diagnosis of Covid-19 infection, but that PCR negativity is of little clinical help, due to his low sensitivity (4). The grouping of patients that have answered the questionnaire based on the decision tree criteria is reported in Fig.4b, among with the results of PCR in each group. Anosmia or ageusia (observed in 45% of tested patients) are highly predictive of PCR+ (86% PCR+), which justifies establishing a clinical diagnosis of Covid-19 infection without performing a PCR. In the absence of ageusia/anosmia, the association of fever and cough (19% of patients) is not specific to Covid-19 infection: PCR is useful in this case (65% of the group is PCR+).

**Figure 4:**
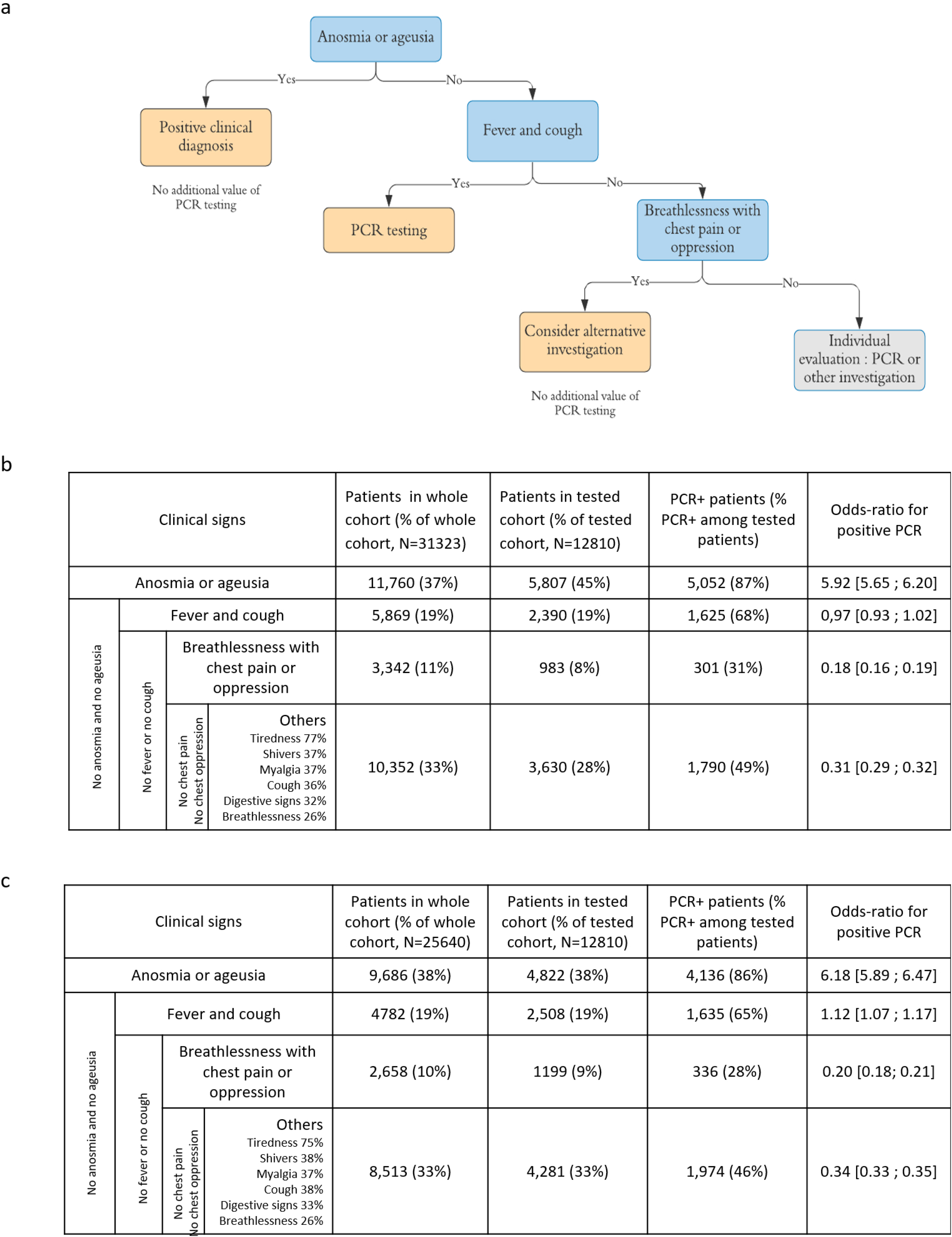
Added value of PCR testing in patients with clinically suspected moderate Covid-19 infection based on their symptoms. a) Proposal of a diagnostic strategy in medical practice, for patients with clinical suspicion of moderate Covid-19 infection by a physician. Anosmia or ageusia are predictive of PCR+ and these patients could be diagnosed based on clinical symptoms only. In case of fever and cough, PCR testing has a good diagnostic value. Then, breathlessness associated with chest pain or oppression is predictive of PCR-, although these symptoms may be signs of Covid-19 pulmonary infection. Therefore, PCR is little valuable in this group and an alternative investigation, such as chest imaging, is indicated. For the remaining patients, PCR is of little added value, but this group is heterogeneous and PCR or alternative investigations should be considered depending on the clinical presentation. b) Description of the different clinical groups defined by the decision tree. The non-weighted numbers, proportions and odds-ratios of being PCR+ in each group are provided. PCR testing has a different added value for each group: it is less useful for groups with PCR+ odds ratios far away from 1. c) Same description as Fig. 4b) with propensity-score weighting. Findings are similar.

Cardiopulmonary symptoms, i.e. breathlessness with chest oppression or pain, are predictive of negative PCR results (28% of the group is PCR+, 8% of tested patients). PCR thus has a poor diagnostic value in this case, justifying the use of another diagnostic investigation, especially in those patients who may be at high risk of complication. The rest of the patients is a heterogeneous group presenting with flu-like illness, moderate breathlessness or digestive symptoms and could benefit from PCR or other investigations depending on the physician evaluation.

Fig.4c reports numbers without propensity-score weighting, with similar findings. Among patients that have answered the survey, the decision tree validates the diagnosis of Covid-19 without any investigation in 11,760 patients (true counts, 38% of the whole cohort). It leads to maintaining PCR as a first-line diagnostic tool in 19% of cases. Among patients without anosmia/ageusia and with cardiopulmonary signs, chest imaging was only performed in 18% (13% among the complementary group), where the decision tree recommends systematic testing.

## Discussion

In this study, we estimate a model that predicts PCR results from clinical presentation, based on 12,810 symptomatic patients with suspected Covid-19 infection, in order to prioritize PCR access. Until now, several tests have been used to confirm the diagnosis of Covid-19 infection, and PCR is the closest to a gold standard. It is a very specific test for this disease, with a high positive predictive value. Yet it is not sensitive to the disease in general (4), due to the possible presence of virus in other localizations and to many biases in its realization (9). High priority patients who should be tested include hospitalized patients, symptomatic healthcare workers, symptomatic residents in congregate living settings, and selected contacts. Ideally, all symptomatic patients, regardless of the symptoms, and all selected asymptomatic people should be tested (10). Other investigations, especially chest imaging, improve the sensitivity of Covid-19 diagnosis in some groups of patients (11). Serological tests are developing but turn positive late for diagnosis (12). In this context, a diagnostic strategy combining clinical evaluation and targeted investigations is necessary, to ensure effective identification of all cases and security of patients at risk of severe disease. Artificial intelligence has been used to optimize diagnostic testing in the epidemics context, based on CT-scan automatized interpretation, whereas our approach is based on the clinical presentation (13).

The COVIDOM cohort is, to the best of our knowledge, the largest ambulatory population with suspected Covid-19. Patients are younger than in the series of hospitalized patients and predominantly women (2). Our clinical findings are congruent with data already published. Focusing on the patients with chemosensory symptoms, who form an important clinical subgroup in our decision algorithm, we found the same characteristics in the COVIDOM cohort as in the literature: in comparison with Covid-19 patients in general, they are younger, predominantly women, and they have fewer comorbidities, except for asthma which was more frequent (14,15). According to recent discussions, anosmia is likely due to both mucosal inflammatory reaction and central nervous system infection through the olfactory nerve (22), as described for SARS-CoV in mice (23).

Access to PCR in the COVIDOM cohort does not follow the theoretical guidelines: PCR is neither systematically performed on hospitalized patients (80%), nor on symptomatic patients (41%). Some groups of patients, not supposed to be at high risk, including women and non-smokers, are tested more often (both 43%). In addition to showing that PCR is not available for all patients who should be tested, Fig.1c underlines that some patients with *a priori* higher respiratory risk (any comorbidity, smokers) are less likely to be tested (p<0.0001), which was not expected. This disparity in PCR testing is not due to explicit medical targeting, but to limited access, which confirms the need for a strategy to prioritize those tests. Although political decisions are being taken to make testing as widely available as possible, the number of PCR will be limited. Moreover, performing PCR in some patients will not systematically help us to correctly diagnose, isolate and treat.

We found that, in the Covid-19 epidemic context, the clinical presentation is predictive of the positivity of PCR in some groups. Patients with anosmia or ageusia represent 50% of our cohort, 47% of symptomatic patients with PCR+ in Europe (14), and up to 86% of symptomatic patients referred to an ENT clinic (15). We show that 90% of tested patients with these symptoms (OR 5.61-6.14) turn out to be PCR+, which is consistent with the high positive predictive values of anosmia (84.7%) and ageusia (88.1%) for SARS-CoV-2 infection found by Fontanet et al. also in France (16). In the current context, those patients can be considered as infected with Covid-19 with little approximation even without PCR testing. This validates with statistical rigor the empirical recommendations found in the literature (17,18). In addition, the specific form that these symptoms take for Covid-19 limits the risk of false positive diagnosis: although the prevalence of olfactory and taste dysfunction in adults ranges between 3.8% and 13.5% (19,20), with 39% of cases retrospectively attributed to an upper respiratory tract infection (21), unexplained sudden onset anosmia or ageusia is extremely rare in clinical practice. Finally, our analysis is based on a very coarse evaluation of anosmia or ageusia, defined as the recent onset of loss of smell or taste, as reported by the patient in a multiple choice question. There is no doubt that critical and precise medical history taking helps reduce the rate of false positive diagnoses (10% of PCR-in our cohort, which means maximum 10% of differential diagnosis), thanks to the descriptions of SARS-CoV2-associated olfactory and gustatory symptoms published recently (14,15).

Our analysis then underlines that in patients without chemosensory symptoms but with cough and fever, which are non specific symptoms, representing 19% of our cohort, PCR results cannot be predicted and testing has a relevant clinical value. PCR has a useful positive value and seems clinically relevant to separate differential diagnoses, although PCR+ is not systematically evidence of Covid-19 and proves only SARS-Cov-2 presence in the sample (4).

Finally, our study singles out a third particular group, namely patients with breathlessness and chest pain or oppression, among those without anosmia/ageusia and fever with cough, namely 11% of our cohort. PCR testing has a poor diagnostic value in this group, with only 30% of positive results. Whether the remaining patients have Covid-19 with PCR- or a differential diagnosis is not established in our series. In both situations, chest CT-scan, whenever available, may be more reliable by showing specific pulmonary lesions, such as ground-glass opacities (13). One physiopathological hypothesis in Covid-19 transmission is that the SRAS-CoV-2 is transmitted through the upper respiratory tract, where it could either remain, after a general efficacious inflammatory reaction, or spread to the lower respiratory tract, causing severe pneumonia. In this scenario, anosmia and ageusia would not only be of high diagnostic value: they may be predictors of good outcome (19). This finding is compatible with the findings in our cohort: the non anosmic/ageusic group is more likely to be admitted in hospital. A positive PCR thus appears to be an indicator of persistent virus in the nose and throat, whereas a negative PCR may indicate migration of the virus and potential respiratory complications.

In the COVIDOM cohort, applying the diagnostic strategy would lead to making a purely clinical diagnosis in 37% of symptomatic cases. This would allow better targeting of PCR testing, particularly indicated in non anosmic/ageusic patients with fever and cough. The other patients who additionally present breathlessness, chest pain or oppression will rather benefit from another first-line investigation: 70% of PCR are negative in this case despite Covid-19 suspicion, which suggests a high false negative rate.

As PCR testing has a low sensitivity to Covid-19 infection,we may wonder how many times this test should be replicated for reliability. In some studies, PCR testing was performed as many times as necessary to confirm the infection when clinical suspicion was very high (22,23). In our study, the mean number of tests was 1.20 (SD 0.64) and did not differ in particular population groups. The clinical presentation would be a clue for the clinician to decide whether repeating the test will be useful, and the evolution may also lead to alternative investigations. Moreover, as already observed (10), the delay between the first symptoms and the PCR test has a significant effect on the PCR result (odds of being PCR+ are 1.03-1.04 times lower on day J+1 than day J, cf Fig.S3b), which prompts to perform PCR as early as possible, in the absence of chemosensory symptoms.

There are a number of important limits in our study. First, the results of this study are to be interpreted in the specific setting of the Covid-19 epidemics. The cohort is recruited in a region with a high prevalence of the disease, with up to 26% of infected people in some areas of France (16). The criterion motivating inclusion in the COVIDOM cohort is the clinical suspicion by a physician of Covid-19 infection. This inclusion criterion is subjective, and highly depends on the epidemiological context in the area. Some patients with a differential diagnosis have inevitably been included in the cohort. PCR-may be due, in our series and in general practice, to a false negative result or to a differential diagnosis, so that PCR-is actually considered as diagnostically inconclusive, due to its low sensitivity (4).

Most parameters analyzed were harbored from self-reported questionnaires, with an inevitable rate of mistakes. In particular, performance of the test partially relies on the patient’s recollection, whereas PCR results are verified by a medical database. In addition, our study falls short of considering that symptoms appear progressively. We analyze symptoms present during the course of the disease; those may not be present during the first medical evaluation. However, anosmia/ageusia are early signs, developing on average 4.4 days after infection (14). They are likely to have developed when the patient seeks medical care (median 5 days, IQ2-8, after the beginning of the symptoms in the COVIDOM cohort). Less importantly, we only analyze the symptoms provided by the forms submitted to the patients. These forms did not investigate all possible clinical signs; additional symptoms were identified manually in the patients commentaries, especially headaches, vertigo and upper respiratory tract symptoms. Preliminary analyses show that these items are not key symptoms to predict PCR results; yet they should be probed in future clinical investigations.

Finally, in the methods we used, our analysis of PCR results is based on the tested population, which is not representative of all patients infected with SARS-CoV-2. We challenged our reweighting strategy by reproducing the same analysis without this strategy and note that a similar decision tree is obtained, which shows that the final diagnostic algorithm is reliable. It could also be completed by adding the date of the beginning of the symptoms and the patient’s characteristics as predictors, in addition to the clinical presentation.

## Conclusion

In the context of SARS-CoV-2 epidemics, we found that the clinical presentation is predictive of PCR+ in anosmic/ageusic patients, allowing to make the diagnosis without any investigation. We propose that patients with fever and cough be tested by PCR, and we show that PCR does not provide useful results for patients with breathlessness and chest pain or oppression. In that group, chest imaging as a first-line investigation may be more useful. Our findings will help target PCR tests in the symptomatic population, and contribute to crafting the best strategy to manage the pandemic.

## Data Availability

Data and code available on request for academic researchers

## Acknowledgments

Data used in preparation of this article were obtained from the AP-HP Covid CDW Initiative (ACCI) database. A complete listing of the ACCI members can be found at: (https://eds.aphp.fr/covid-19).

We thank FALZON Alexandre, FAYOLLE Guillaume, LAPORTE Fanny, Amélie TORTEL and all the Nouveal-e Santé team for their help in the web application and regional center surveillance interface development. We also thank DEBASTARD Laurent, GRENIER Alexandre, HODY Julien, PENN Thomas and the Paris region URPS (Union régionale des professionnels de santé) for their help in the development and spreading of the Covidom solution.

We thank the Polytechnique network for helping with the volunteers recruitment.

## Authors Contributions

APRA Caroline, CAUCHETEUX Charlotte and MENSCH Arthur were involved in the study conception, data extraction, data analysis, interpretation of results, drafting the manuscript and approving the final version of the manuscript.

MANSOUR Jenny was involved in the study conception, data extraction, data analysis, and approving the final version of the manuscript.

BERNAUX Mélodie was involved in the study conception, data extraction, data analysis, interpretation of results, and approving the final version of the manuscript.

DECHARTRES Agnès was involved in the study conception, data analysis, interpretation of results, critically revising the manuscript and approving the final version of the manuscript.

DEBUC Erwan was involved in critically revising the manuscript and approving the final version of the manuscript.

DINH Aurélien was involved in interpretation of the results, critically revising the manuscript and approving the final version of the manuscript.

LESCURE Xavier was involved in COVIDOM development, in interpretation of the results, critically revising the manuscript and approving the final version of the manuscript.

YORDANOV Youri was involved in the study conception, data analysis, interpretation of results, critically revising the manuscript and approving the final version of the manuscript.

JOURDAIN Patrick was involved in COVIDOM development, in the study conception, interpretation of results, critically revising the manuscript and approving the final version of the manuscript.

GRAMFORT Alexandre was involved in the study conception, data extraction, data analysis, interpretation of the results and approving the final version of the manuscript.

PARIS Nicolas was involved in EDS data access and function, and approving the final version of the manuscript.

## Ethical approval

This study received the ethical approval of the ethical committee of AP-HP (IRB00011591).

## Data sharing

Data available upon request for academic researchers.

